# Somatic mutations reveal clonal cell populations in atherosclerotic plaques

**DOI:** 10.1101/2022.05.16.22275001

**Authors:** LB Steffensen, S Kavan, PS Jensen, S Møller-Larsen, MK Larsen, M Dembic, LvB Andersen, JS Lindholt, KC Houlind, M Thomassen, LM Rasmussen

## Abstract

**Background:** In recent years, the potential involvement of clonal cell populations in atherosclerosis has gained renewed interest. Evidence from independent research groups unambiguously showed that smooth muscle cells clonally expand in experimental atherosclerosis, and clonal hematopoiesis of indeterminate potential (CHIP) has been proposed as a novel risk factor for atherosclerotic cardiovascular disease in humans. However, direct evidence for the presence of clonal cell populations within human atherosclerotic tissue is lacking.

Here, we investigated whether characterizing the mutational landscape of human atherosclerotic plaques could provide insights into the role of clonal cell populations in human atherosclerosis.

**Methods and Results:** We conducted *in-depth* whole-exome sequencing of 65 samples derived from 13 carotid plaques obtained from patients undergoing carotid endarterectomy. This approach unveiled a landscape of somatic mutations exclusively confined to plaque tissue (*i.e.*, not detected in patient-matched buffy coats). Leveraging variant allele frequencies, we inferred that distinct locally expanded clones often accounted for over 10% of the cell population within the plaque samples, and certain clones extended across multiple segments within the same plaque, necessitating a physical clone size of at least 1 cm. Furthermore, clones carrying multiple mutations spanned diverse plaque regions, encompassing the media, sub-core intima, and necrotic core. We did not detect any mutational hotspots, however, 21 of the mutated genes appeared in the Integrative Onco Genomics database of 619 mutational cancer driver genes (χ^2^ test: *p* = 0.015). Moreover, unbiased pathway enrichment analysis disclosed enrichment of terms linked to cell-cell adhesion and the contractile apparatus, and genes underlying these enrichments were primarily expressed by smooth muscle cells, endothelial cells, and fibroblasts, as corroborated by analysis of single-cell RNA sequencing data. Finally, six patients were CHIP carriers, and in several of these patients, hematopoietic clones constributed significantly to the cell population of plaque segments.

**Conclusions:** In summary, our study provides compelling evidence that somatic mutations are an intrinsic aspect of human plaques. Moreover, we substantiate that specific mutation-carrying cells within plaque tissue undergo expansion, giving rise to clones of substantial size. We propose that unraveling the fundamental underpinnings of clonal biology and the phenotypic traits of expanding clones may unveil novel mechanisms to impede lesion development or enhance plaque stability.

## INTRODUCTION

Atherosclerosis is the focal accumulation of lipid, fibrous tissue, and cells in the intimal layer of arteries. The disease develops over decades and may suddenly manifest as myocardial infarction or ischemic stroke - leading causes of death in the world^1^.

The cellular component of atherosclerotic plaques mainly originates from circulating myeloid cells and smooth muscle cells (SMCs) recruited from the local medial layer^2^. Recent advances in the field have revised our perception of both sources^3^: First, by use of multicolor lineage tracing in mouse models, it was unambiguously demonstrated that all SMC-derived plaque cells are progeny of few medial SMCs^4,5^, thus reviving the hypothesis of monoclonality in atherosclerosis proposed by Benditt and Benditt almost 50 years ago^6^.

Second, several lines of evidence now support an important role for clonal hematopoiesis of indeterminate potential (CHIP) in atherosclerotic cardiovascular disease^7–9^. Individuals with CHIP carry expanded hematopoietic clones often harboring mutations in epigenetic regulators (*e.g.*, *DNMT3A* and *TET2*)^10^, which besides conferring a selective advantage render myeloid cells more inflammatory and pro-atherogenic^11^.

Although these recent advances point to the involvement of clonal cell populations in the pathology of atherosclerosis, the presence and the extend of clonal populations partaking in human atherosclerosis remains elusive.

Recent studies have demonstrated that somatic mutations and clonal cell populations are present in a range of non-tumor tissues besides CHIP^12^ (*e.g.*, skin^13^, prostate^14^, bladder^15^, and esophagus^16^), and in various pathological conditions (*e.g.*, chronic liver disease^17^, inflammatory bowel disease^18^, and endometriosis^19^). There are indications that mutation-carrying clones partake in disease pathophysiology, and moreover, the burden of somatic mutations and clone sizes have been shown to aggravate with age and proposed to be involved in age-associated reduction of organ function^16,20^.

In this study, we hypothesized that similar processes take place in human atherosclerotic plaques, and that interrogating the somatic mutational landscape of plaques may provide unprecedented insight into the potential involvement of clonal cell populations in atherosclerosis. Identification of mutations in single or few cells is challenging, but somatic mutations amplified by proliferative expansion of mutation-carrying cells can be detected by whole-exome sequencing of bulk tissue samples as recently shown in the context of endometriosis^19^.

To address our hypothesis, we performed deep whole-exome sequencing of human atherosclerotic plaques and simultaneously sequenced buffy coat DNA from the same patients to evaluate whether detected mutations were confined to plaques (*i.e.*, not detected in buffy coats). In addition, parallel sequencing of plaque tissue and buffy coats from the same individuals enabled us to investigate the contribution of circulating CHIP mutation-carrying cells in the plaque cell population.

## RESULTS

### Somatic mutations and locally expanded clonal cell populations are inherent features of atherosclerosis

Whole-exome sequencing was performed on DNA extracted from carotid plaques and patient-matched buffy coats from 13 patients undergoing carotid endarterectomy (**supplementary table S1**). From each carotid plaque, one to four segments (2-5 mm tissue per segment) distanced 4-10 mm were analyzed in parallel (**fig. 1a**). A subset of plaque segments were further subdivided while preserving morphological context of sequenced samples (**fig. 1b**). Every sample underwent two rounds of independent sequencing starting with raw DNA. The median sequencing depth across samples was 700X (IQR: 518-932.5) with an inferred base call accuracy of 99.9%, enabling accurate identification of mutations at variant allele frequencies (VAF) ≥ 1% with a variant read depth ≥ 5 (median: 17 (IQR: 12-26)) (all sequencing metrics are available in **supplementary table S2**).

**Figure 1.**
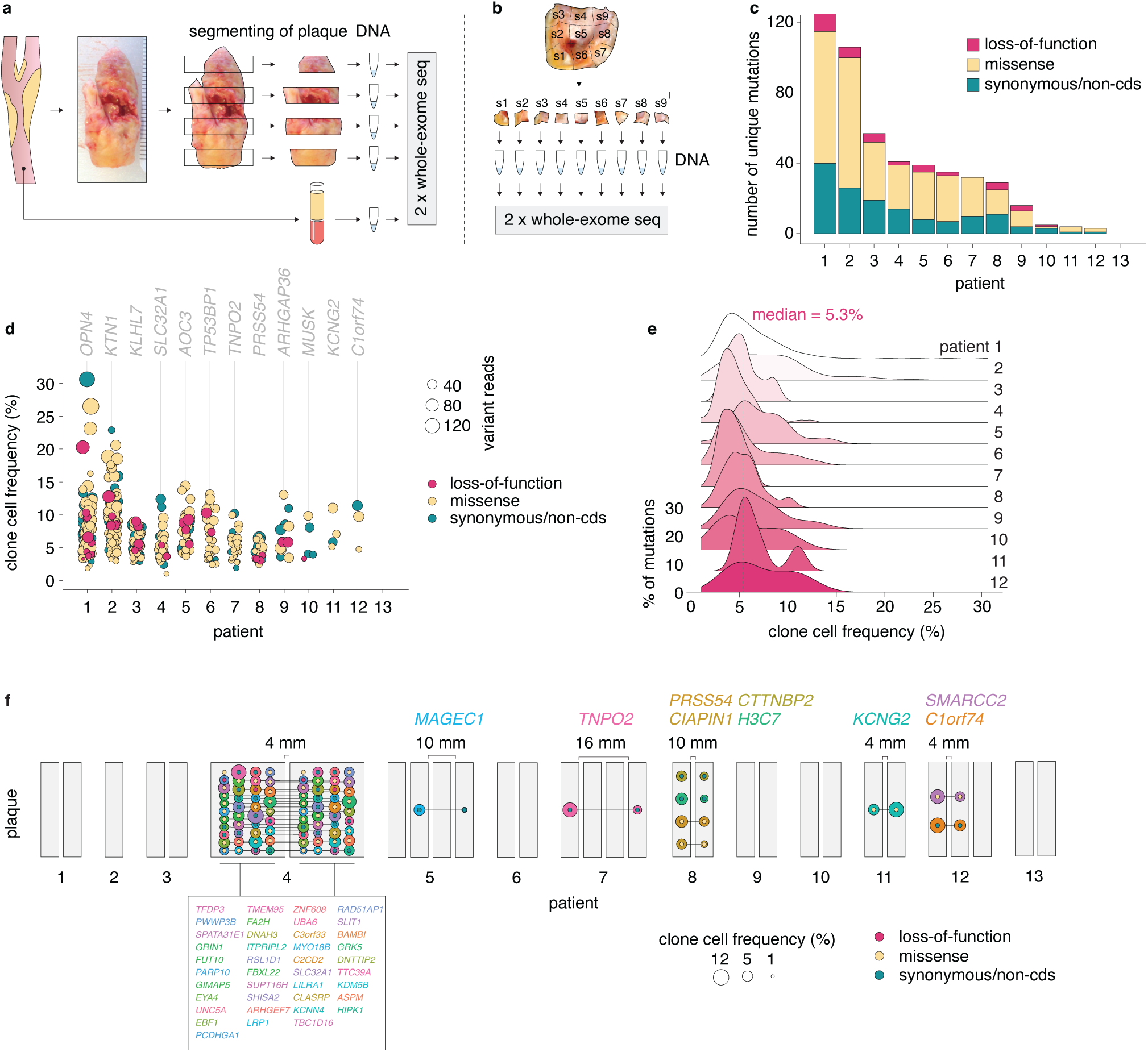
Somatic mutations and locally expanded clonal cell populations are inherent features of atherosclerosis. **a-b.** Carotid atherosclerotic plaques (one to four segments per plaque) and buffy coats from 13 patients undergoing carotid endarterectomy were analyzed by whole-exome sequencing (a). Four segments were further subdivided for sequencing while preserving morphological context (b). Every sample underwent two rounds of independent sequencing, starting with raw DNA. **c.** Barplot showing the number of plaque-confined somatic mutations (*i.e.*, mutations not identified in patient-matched buffy coats) for each patient. Mutation effect is indicated by color. non-cds refers to mutations present in introns or intergenic regions. **d.** Frequency of clone cells (calculated from variant allele frequencies, VAFs) carrying a specific somatic mutation in plaque samples for each patient. Although single clones may harbor several mutations, each mutation is represented by separate clone cell frequencies on the plot. Mutation effect is indicated by dot color, and variant reads is indicated by dot size. For each patient, the mutated gene representing the highest clone cell frequency is shown. **e.** Density plot showing the distribution of clone cell frequencies for each patient (except for patient 13 where no mutations were detected). The area under each curve corresponds to 100% for each patient. **f.** In several patients, somatic mutations were detected in more than one plaque segment from the same patient. Plaque segments are represented by grey bars, and mutations shared by more than one segment are shown as interconnected colored dots. Dot colors correspond to the shown gene symbols and dots sizes show clone cell frequency. The mutation effect is indicated by the smaller dots within mutation dots. Intersegment distances used to estimate physical extends of clonal populations are shown.

With these criteria, we identified on average 37.8 (range: 0-125, median: 33.5, IQR: 13.25-45) somatic mutations per plaque (**fig. 1c**). These mutations were not detected in patient-matched buffy coats supporting local (within plaque tissue) amplification of mutations by expansion of mutated cells (*i.e.*, clones) by proliferation. Clone cell frequencies (calculated from VAFs) ranged from 1% (the pre-defined limit of detection) to 30.6% (median: 5.3%, IQR: 3.9-7.7%) (**fig. 1d-e** and **supplementary table S3**).

Somatic mutations were detected in 61 of the 65 analysed plaque samples ranging from 1-80 unique mutations per sample. For several plaques, specific mutations were identified in more than one segment, requiring a physical extend of clones harboring these mutations of at least 4-16 mm (**fig. 1f**).

To evaluate the number of cells constituting individual clones, we calculated the number of sequenced cells from plaque segment DNA yields, assuming 6 pg DNA per cell. Sample cell numbers ranged from 2,150 to 1,950,000 (median: 261,667, IQR: 133,833-717,500) resulting in cell number per clone estimates of 198 to 139,159 (median: 9,186, IQR: 4,636-22,390). Since DNA purifications may be sub-efficient, these are conservative lower-bound estimates.

### Pathway enrichment suggests role of mutations in driving clonal expansion

After establishing the existence of locally expanded mutated clones, we sought to evaluate whether somatic mutations were clone *drivers*, *i.e.*, conferring a selective advantage (analogous to neoplasms), or merely amplified by coincidence as *passengers* in a clone expanding by other mechanisms.

We did not detect any mutational hotspots (as in CHIP^10^ and certain types of cancer^21^), however, 21 of the mutated genes appeared in the Integrative Onco Genomics database of 619 mutational cancer driver genes^21^ (χ^2^ test: *p* = 0.015).

Over-representation analysis of all 463 mutated genes identified across the 13 patients resulted in significant enrichment of terms associated with cell-cell adhesion and the contractile apparatus. This enrichment was primarily driven by protein sequence altering mutations, as repeating the analysis based on the 334 missense and loss-of-function mutations yielded similar results (**fig. 2a**). In contrast, pathway analysis based on the 137 synonymous mutations did not result in pathway enrichment.

**Figure 2.**
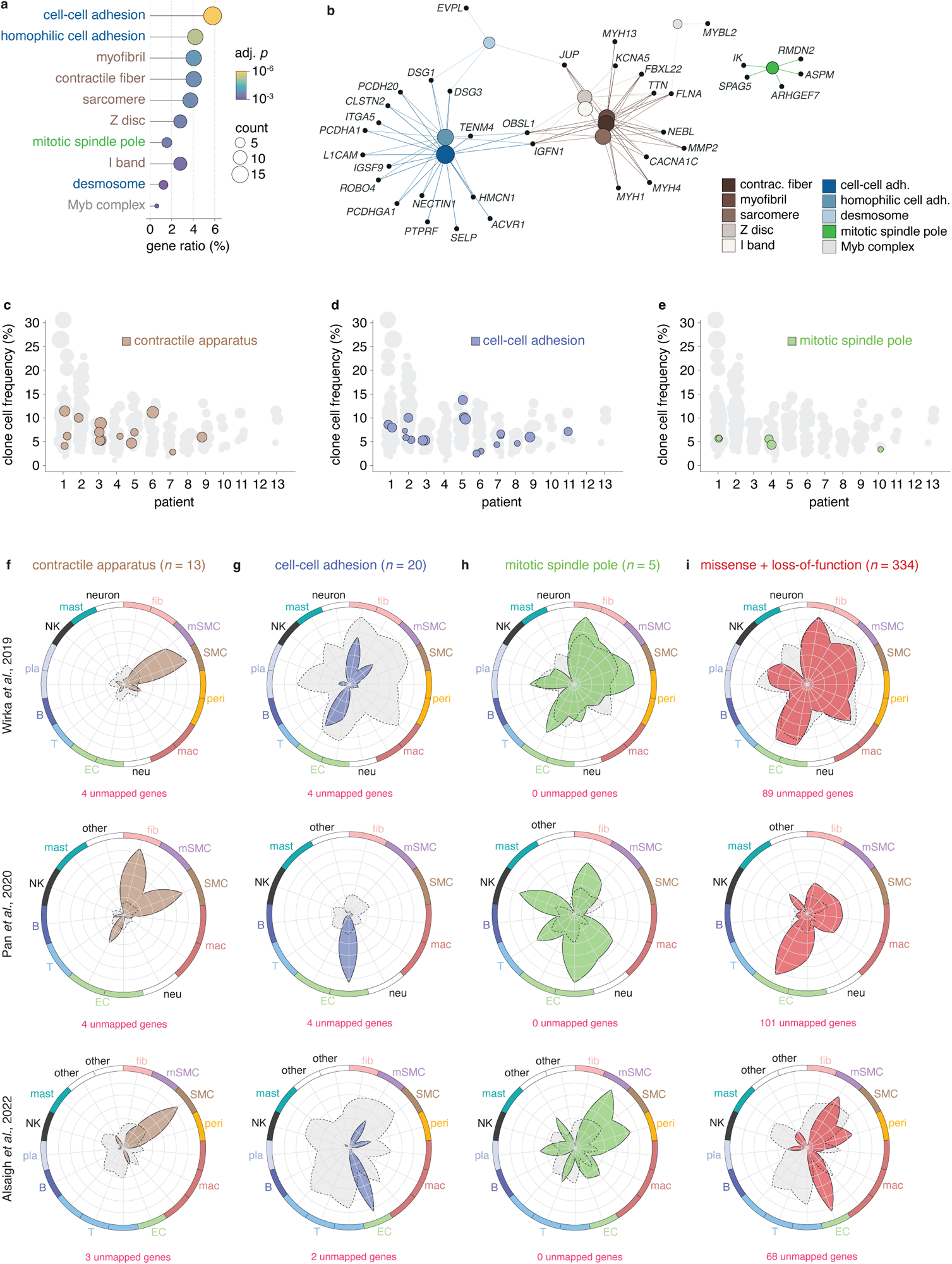
Pathway enrichment suggests role of mutations in driving clonal expansion. **a.** Dotplot showing the enriched gene ontology terms of the 334 genes having a loss-of-function or missense mutation across all patients. Pathway analysis based on the 137 synonymous mutations did not result in pathway enrichment. Dot sizes indicate number of mutated genes for each term. The ratio shows the coverage of a given term by mutated genes, and dot colors indicate significance level of the enrichment. **b.** Network plot showing enriched gene ontology terms and the mutated genes belonging to each term. Enriched pathways could be grouped into three main clusters with minimal overlap: 1) cell-cell adhesion, 2) contractile apparatus, and 3) mitotic spindle pole. **c-d.** Distribution of genes underlying enriched terms across patients. **f-i.** To evaluate the expression pattern of mutated genes, three single cell RNA sequencing datasets of human plaques (Wirka *et al.* 2019^22^, Pan *et al.* 2020^23^, and Alsaigh *et al*. 2022^24^) were used. The mean expression level of loss-of-function and missense mutations were plotted for each cell type in every dataset. This was done for mutated gene belonging to the three main pathway clusters identified in b: contractile apparatus (f), cell-cell adhesion (g), mitotic spindle pole (h), and all loss-of-function and missense mutations identified across patients (i). As a reference, the mean expression level of all genes of the atherosclerosis transcriptome was plotted for each dataset in grey for comparison. The number of unmapped genes, *i.e.*, mutated genes which was not identified in a given single cell RNA sequencing dataset, is shown below each plot. fib = fibroblast, mSMC = modulated SMC, peri = pericyte, mac = macrophage, neu = neutrophil, EC = endothelial cell, T = T cell, B = B cell, pla = plasma cell, NK = natural killer cell, mast = mast cell, other = un-annotated clusters in original publication.

Enriched pathways could be grouped into three main clusters: 1) cell-cell adhesion, 2) contractile apparatus, and 3) mitotic spindle pole (**fig. 2b**). The genes underlying these enrichments were found distributed among a broader group of patients instead of being confined to just a few individuals (**fig. 2c-e**). To investigate which cell populations in atherosclerotic lesions that express the genes driving these enrichments, we leveraged three single-cell RNA sequencing datasets of human plaques^22–24^. The mean expression of mutated genes associated with the contractile apparatus (*n* = 13) was markedly higher in SMC, modulated SMC, and fibroblast cell clusters (**fig. 2f**). Conversely, mutated genes associated with cell-cell adhesion (*n* = 20) exhibited high expression levels in endothelial cells, as well as in other resident vascular cell types, such as fibroblasts, modulated SMCs, and pericytes, across all three datasets (**fig. 2g**). The consistency of expression for mutated genes linked to the mitotic spindle pole across the three datasets was lower, potentially due to increased variation resulting from the limited number of genes (*n* = 5) (**fig. 2h**).

Overall, the expression pattern of missense and loss-of-function mutated genes resembled that of the background atherosclerosis transcriptome in the dataset from Wirka *et al.*. For the two other datasets, however, mutated gene expression showed a notable bias towards resident vascular cell types and away from immune cells (*e.g.*, macrophages and lymphocytes) (**fig. 2i**). These patterns suggest that the identified mutations, if functionally meaningfull, are likely to have a greater impact on resident vascular cells compared to immune cells.

Taken together, although we did not identify general hotspot clonal driver genes with the same gene mutated in several plaques, our analyses indicated a non-random distribution of mutations throughout the exome. Although this implies that some somatic mutations contribute to clonal expansion, further experimentation is required to draw such conclusions.

### Mutated clones span several regions of the plaque

Four plaque segments from different patients (patient 1, 2, 4, and 6) (**fig. 3a**) were dissected while preserving morphological context of each of the resulting samples (**fig. 3b_p1,p2,p4,p6_**).

**Figure 3.**
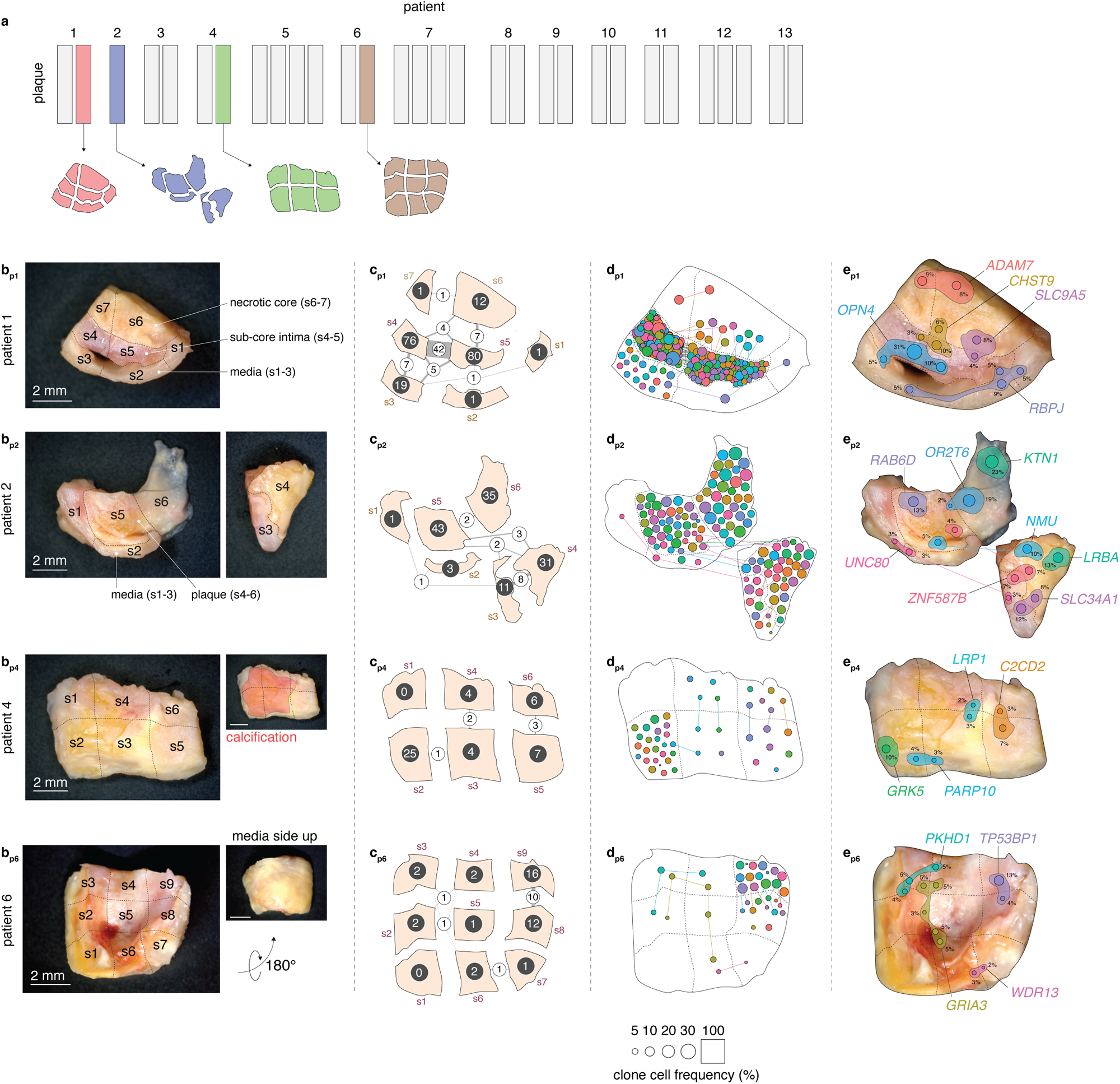
Mutated clones span several regions of the plaque. **a.** To further assess the clonal architecture of atherosclerosis, plaque segments from four patients were subdivided while preserving morphological context. **b.** Depicted are images of the plaque segments, illustrating the division process into distinct samples. In b-e, the subscripts p1, p2, p4, and p6 indicate the patient origin of each segment. **c.** The number of mutations identified in each sample is represented by white numbers, while the number of shared mutations between samples is denoted by black numbers and the thickness of the inter-sample connection lines. **d.** Mutations detected in each sample are shown as dots, with specific mutations shared among samples connected by lines. The size of each dot corresponds to the clone cell fraction it represents, as indicated in the key. Of note, the sample area does not correspond to 100%, as it is increased to fit the dots. **e.** Possible interpretations of the distribution of clones harboring selected mutations are depicted. The dot color and size mirror the key in d. Additionally, a shaded area has been added for which the size corresponds to the clone cell frequency for each sample, setting the sample area to 100%. The genes that are mutated are indicated.

#### Patient 1 and 2

From patients 1 and 2, the medial layer was separated from the overlying plaque tissue, which was further subdivided (**fig. 3b_p1,p2_**). In both patients, the medial layer samples exhibited markedly fewer mutations compared to the overlying plaque samples.

In the plaque segment of patient 1, the medial layer was divided into three samples (s1-3), and the plaque was divided into two sub-core intimal samples (s4-5), and two necrotic core samples (s6-7) (**fig. 3b_p1_**). Two samples from the medial layer (s1-2) had only one mutation (in *RBPJ, RBPJ^mut^*), which extended throughout all three media samples (s1-3) and protruded into intima sample s5 (**fig. 3c-d_p1_**) (a possible interpretation of the *RBPJ^mut^* clone is depicted in **fig. 3e_p1_**). The third sample from the media (s3) had 19 mutations, 12 of which were also present in the overlying intima sample (s4) (**fig. 3c_p1_**). A subset of these were also present in the neighbouring intima sample (s5). In particular, a mutation in *OPN4* was present in 5% of cells in s3, 31% of cells in s4, and 10% of cells in s5, suggesting the precense of a large media-intima spanning clone (possible interpretation of the *OPN4^mut^* clone is illustrated in **fig. 3e_p1_**).

The two sub-core intima samples (s4-5) exhibited a high abundance of somatic mutations and 51 mutations overlapped between these two samples (**fig. 3c-d_p1_**). From the sub-core intima, 11 mutations extended into the overlying necrotic core (exemplified by *CHST9^mut^* and *SLC9A5^mut^* with possible interpretation of mutation-carrying clones illustrated in **fig. 3e_p1_**).

In contrast to the sub-core region, we only detected a single independent (*i.e.*, not found in other regions) mutation (in *ADAM7*) in the necrotic core, which spanned both necrotic core samples (s6-7) (**fig. 3c-d_p1_**) (possible interpretation of the *ADAM7^mut^* clone illustrated in **fig. 3d_p1_**).

Similar to patient 1, the media samples (s1-3) of patient 2 (**fig. 3b_p2_**) had relative few mutations (**fig. 3c-d_p2_**). A single mutation (in *UNC80*) spanned all media samples, but was not found in any plaque samples (**fig. 3c_p2_-d_p2_**). Media sample s3 had 10 mutations in common with plaque sample s4. The most abundant of these mutations was *ZNF587B^mut^* (which also extended into s5) and *SLC34A1^mut^* (possible interpretation of clones carrying these mutations is illustrated in **fig. 3e_p2_**).

In contrast to the medial layer, numerous mutations were identified in the three plaque samples (s4-6), with modest inter-sample overlap (**fig. 3c-d_p2_**). However, several plaque mutations were of considerable size, *e.g.*, *KTN1^mut^* (s5), *OR2T6^mut^* (spanning s5-6), *RAB6D^mut^* (s6), and *NMU^mut^* (spanning s4-5) with clone cell frequencies of 23%, 19%, 13%, and 10% respectively (possible interpretation of clones carrying these mutations is illustrated in **fig. 3e_p2_**).

#### Patient 4 and 6

The plaque segments of patient 4 and 6 were sectioned by a different strategy, where each resulting sample spanned the entire vessel wall (media and overlying plaque) (**fig. 3b_p4_,_p6_**).

In general, fewer mutations were detected in the plaque segments from these two patients. The largest clone detected in patient 4 was a *GRK5^mut^*-carrying clone constituting 10% of cells in sample s2 (**fig. 3_c__-dp4_**). Other mutations overlapped between neighbouring regions (*e.g.*, *PARP10^mut^* (s2-3), *LRP1^mut^* (spanning s3-4), and *C2CD2^mut^* (s5-6) (possible interpretation of clones carrying these mutations are illustrated in **fig. 3e_p4_**).

In patient 6, we observed a mutation in *GRIA3*, which spanned five samples (s2-6). In this plaque, mutations were more confined to samples s8 and s9, which had 10 overlapping mutations. Examples of other sample-overlapping mutations of patient 6 are *PKHD1^mut^* (s2-4), *WDR13^mut^* (s6-7), and *TP53BP1^mut^* (s8-9) (possible interpretation of clones carrying these mutations is illustrated in **fig. 3e_p6_**).

For several samples, the sum of clone cell frequencies calculated from VAFs of each mutation exceeded 100%. Therefore, at least some clones must carry more than one mutation. While the bulk sequencing analysis presented here does not allow for a direct assessment of the mutational architecture of different clones, we observed distinct patterns shared by groups of mutations suggesting that certain mutations may be carried within the same clone.

In particular, mutations identified in the same samples and exhibiting similar magnitude and inter-sample differences in clone cell frequencies (**fig. 4a_p1-p6_**) are likely carried by the same clone, as proposed in **fig. 4b_p1-p6_**. As an example, it is reasonable to assume that the clone carrying *OPN4^mut^* in patient 1 also carries *POU4F3^mut^*, *ZNF800^mut^*, and *LRP1^mut^*, since these mutations are present in the same samples and show comparable differences in clone cell frequency between samples (**fig. 4b_p1_**).

**Figure 4.**
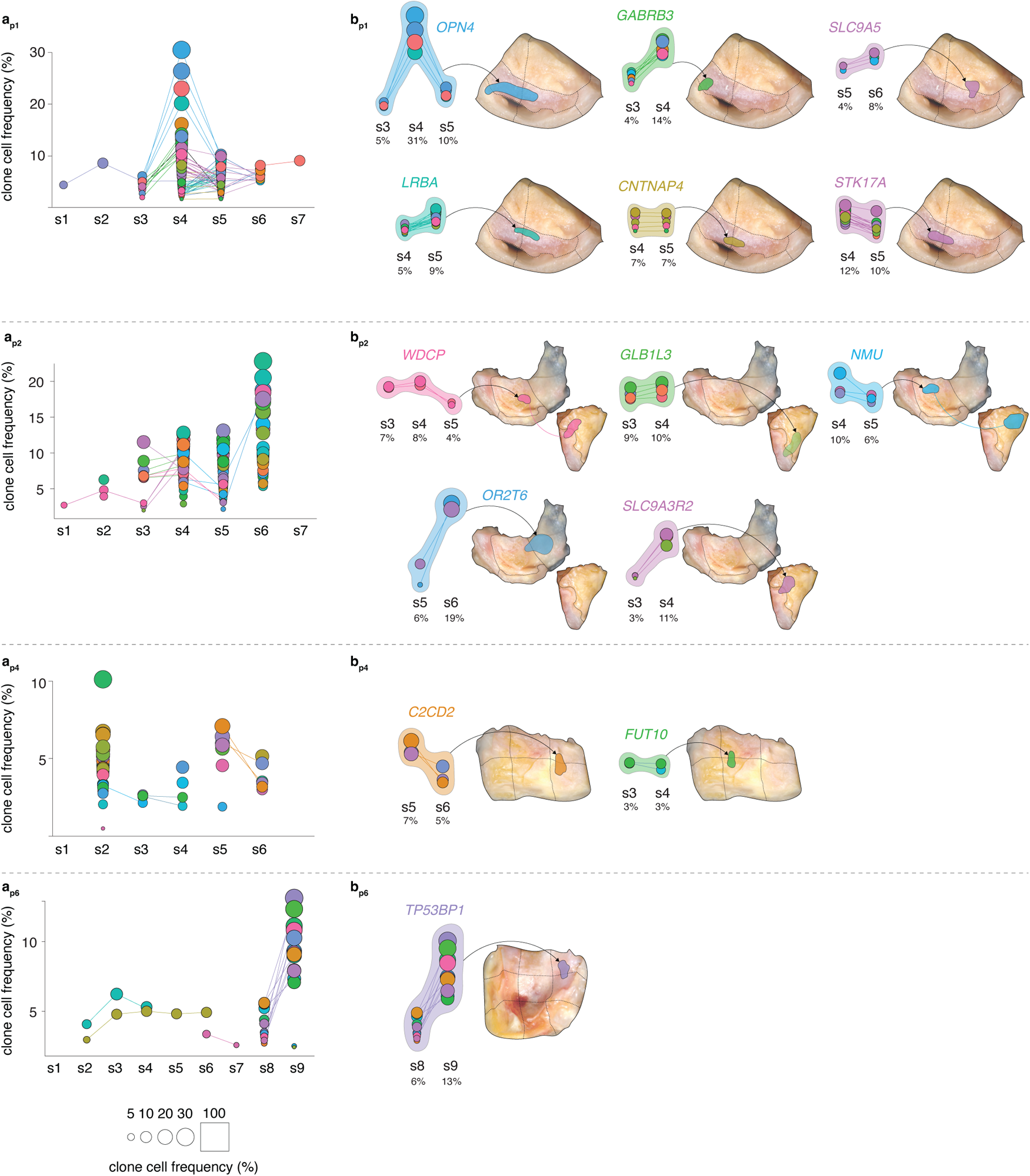
Mutation pattern indicates that some mutations are carried on the same clones. **a.** The plot shows the frequency of clone cells (y-axis and dot size) carrying specific somatic mutations in the same plaque samples as presented in fig. 3. Individual mutations are represented by distinct clone cell frequencies, and lines connect mutations shared among multiple samples. Mutations exhibiting consistent sample presence and parallel trends in clone cell frequency are considered to belong to the same clone. By employing this reasoning, distinct mutation sets displaying similar patterns were identified and presumed to originate from the same clone. These sets are visualized in **b** wherein clone cell frequencies and gene symbols corresponding to the mutations with highest clone cell frequency is shown.

Taken together, although highly descriptive at the level of singe plaques, these data show that locally expanded mutations are present in coherent clones, and that there are regional differences in the burden of mutations, with the necrotic core and medial layer seemed less pronounced, while non-necrotic core plaque area had more.

### Circulating CHIP clones contribute to the plaque cell population

We next investigated the contribution of circulating CHIP mutation-carrying cells to the plaque cell population. By screening for mutations in 78 previously established CHIP genes^10^ using buffy coat DNA, we found 6 of the 13 patients to be CHIP carriers, and several CHIP mutations were detected in each of these patients (**supplementary table S4**). Buffy coat derived VAFs ranged from 1% to 17% (median: 5%, IQR: 2-6%) corresponding to buffy coat clone cell frequencies of 2.1-34.7% (median: 9.7%, IQR: 4.3-11.2%) (**fig. 5a**). In several cases, we identified the same mutations in patient-matched plaque tissue samples. These observations suggest that circulating CHIP-mutated cells infiltrate atherosclerotic plaques, and in some cases constituting a substantial portion of the plaque cell population. For example, in the case of patient 5, a *NOTCH2* loss-of-function mutation had a 22% buffy coat clone cell frequency, and the same mutation was detected in 3 out of 4 plaque samples from the same patient, where it contributed to 13-19% of the plaque sample cell population (**fig. 5b**).

**Figure 5.**
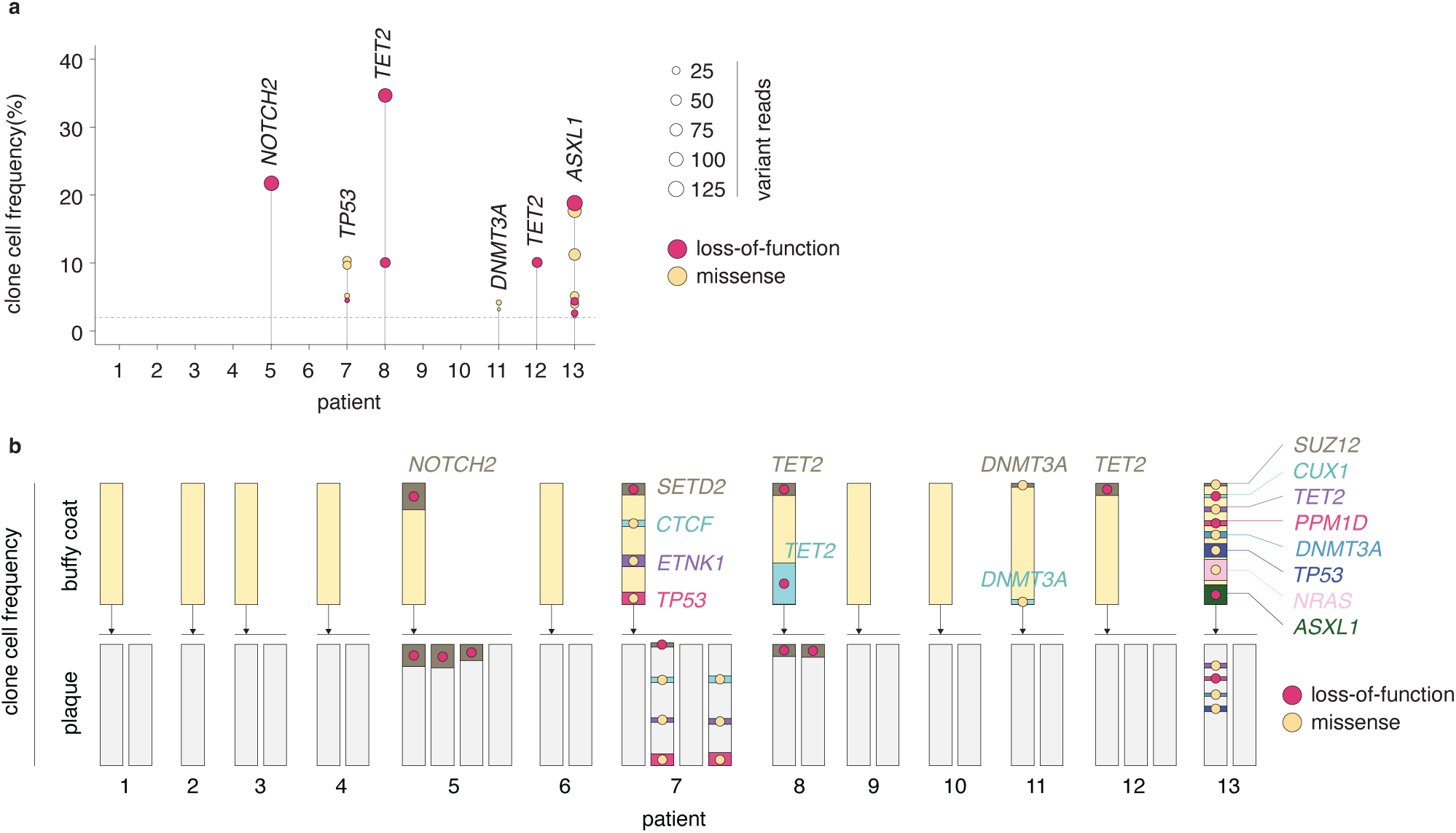
Contribution of clonal hematopoiesis of indeterminate potential (CHIP) clones to the carotid plaque cell population. **a.** Six of the 13 patients were CHIP-carriers. The plot shows clone cell frequencies of CHIP mutations detected in buffy coats. For each patient, the identity of the mutated gene with the highest clone cell frequency is shown. Dot sizes indicate variant reads, and colors indicate the type of mutation. The dashed line shows the defined limit of detection. **b.** CHIP clone cell frequencies in buffy coats (yellow bars) and in plaque segments (grey bars) from the 13 patients. Clone cell frequencies are represented by colored areas within yellow and grey bars. Mutation type is indicated by dot color.

## METHODS

### Ethical approval and patient sample processing

The study was approved by the Danish National Committee on Health Research Ethics (Project ID: CVK-2006749) and The Regional Committees on Health Research Ethics for Southern Denmark (Project ID: S-20140202) and is registered in the Registry of Research Projects in the Region of Southern Denmark (file no. 18/20280).

Carotid plaque tissue and buffy coats from 13 later deceased patients who had undergone carotid endarterectomy either at the Department of Cardiac, Thoracic and Vascular Surgery, Odense University Hospital (OUH), or at the Department of Vascular Surgery, Kolding Hospital, were included in the study. Patients were admitted for surgery because of carotid plaque-associated symptoms. Informed consent was obtained from all participants prior to surgery.

Immediately after surgery, carotid plaque samples were placed and kept in ice-cold Hanks Buffered Salt Solution containing 10 mM HEPES (Biological Industries, SKU BI-02-016-1A and BI-03-025-1B, respectively). Within 24 hours, plaques were sliced into 2 mm segments (patients 5, 7, 11, 12, and 13) or 5 mm thick segments (patients 1, 2, 3, 4, 6, 8, 9, and 10) using a custom-made device. Every third segment was frozen at -80°C and stored in the Odense Artery Biobank infrastructure of the Centre for Individualized Medicine in Arterial Diseases (CIMA) at OUH. Four plaque segments were further subdivided to assess the distribution of mutations while preserving morphological context.

Blood was collected in EDTA vials, centrifuged at 4,000 x g for 10 minutes at 4°C, and buffy coats were collected using a pipette from the top of the centrifuged sample, and stored at -80°C in the Odense Artery Biobank.

### Whole exome sequencing (WES) and data analysis

Whole exome sequencing was performed on blood (buffy coat) and tissue samples obtained from the 13 patients. Genomic DNA was isolated from fresh-frozen tissue samples and buffy coats using standard methods. The libraries were prepared using 50 ng DNA and Twist Comprehensive Exome Panel kit (Twist Bioscience, San Francisco, US), and 2×150bp pair-end sequencing was performed on an Illumina NovaSeq 6000 platform. Unique dual indexing was applied to reduce cross sample contamination from index hopping. Furthermore, unique molecular identifiers (UMI) from IDT were applied for error suppression.

Sequencing data from tissue sampes were processed by Illumina DRAGEN Bio-IT Platform v4.03 (Illumina, San Diego, CA, U.S.) using the DNA pipeline in somatic mode. The raw reads were aligned to Illumina DRAGEN Graph Reference Genome GRCh38. Error corrections were performed by collapsing of PCR duplicated reads by UMI. Buffy coat fastq files were aligned to reference sequence (human NCBI build 38) using the Burrows-Wheeler algorithm (BWA-MEM) version 0.7.12 and processed with the Genome Analysis Toolkit (GATK 4.2.0.0, gatk.broadinstitute.org) to generate recalibrated BAM files. Shortly, files were sorted using SortSam (version 1.3.1) and duplicates removed using MarkDuplicates (Picard Toolkit, version 2.25.0). Base quality score recalibration was done using BaseRecalibrator and ApplyBQSR to output post-recalibration BAM files used in the analysis. Raw data quality check was done using FastQC (version 0.11.9). After data processing, Qualimap (version 2.2.2) and Picard were used to check mapping quality. Information gathered from Qualimap is shown regarding GC content, insert size, coverage, percentage of aligned bases, and general error rate. Information about mapping, and duplication rate was gathered using Picard.

Somatic mutations are called by finding mismatches present in a sample of interest and a matched normal sample (buffy coat from the same patient). We sequenced both matched buffy coat and tissue samples at high coverage (median: 700X (IQR: 518-932.5)) to be able to identify with high confidence somatic variants and distinguish them from sequencing artifacts, even at lower VAFs. We set the lowest VAF at 1%, and discarded variants with under 5 reads. Mutations identified in two independent sequencing rounds (starting from raw DNA sample) from the same patient were included in the subsequent analyses. The mutations presented in the results are those that were consistently identified in more than one independent sequencing for each plaque. To determine the clone cell frequencies, we calculated them from VAFs obtained in both sequencing rounds, while taking into account the number of reads per sequencing round. Clone cell frequencies were calculated based on the assumption of mutation heterozygosity by multiplying VAFs by 2, while VAFs of mutations located on the X-chromosome in male patients was multiplied by 1.

To identify somatic variants occurring in the tissue samples and remove germline SNPs, Mutect2 joint variant-calling was performed on matched samples tissue vs. buffy coat. For very high sensitivity calling of variants identified in one sample, MuTect force calling was performed on all other samples. For the identification of single nucleotide variants and indels occurring in both buffy coat and frozen tissue, we used MuTect2 in single sample mode. Common single-nucleotide polymorphisms (SNPs) were removed by filtering against a panel of normal generated by the 1000 Genomes Project samples, and by using population allele frequencies of common and rare variants from the Genome Aggregation Database (gnomAD) (https://console.cloud.google.com/storage/browser/gatk-best-practices/). Variants were annotated using VarSeq version 2.2.1 (Golden Helix, Inc.) and filtered through a cascade of filters.

Briefly, for variant calling in non-matched samples, the filters were: (i) variant is in a CHIP gene according to our list of 78 CHIP genes^10^, (ii) the region is covered by at least 20 reads, (iii) has a known or predicted damaging effect on protein functionality (according to ClinVar or prediction tools), and (iv) has a minor allele frequency (MAF) < 1% (according to gnomAD). gnomAD is the world largest public collection of genetic variation, and it is based on 125,748 exomes and 15,708 whole-genomes collected worldwide (https://gnomad.broadinstitute.org/). We used the gnomAD collection from exome and genome sequencing data as a large external control group. For matched MuTect2 calling tissue vs. buffy coat, we selected variants with (i) at least 5 reads, (ii) a VAF in reads ≥ 1%, and no or very low signal in the buffy coat (< 0.01%), to eliminate variants that are present also in the buffy coat and were erroneously included by the variant calling process. Built-in post-calling filters in MuTect2 were used to select for high quality variants. A base quality threshold of 25 was set to accept a variant. All filtered variants were inspected manually using VarSeq; mapping artifacts and low-quality calls were removed.

For functional prediction of nonsynonymous variants we used the Combined Annotation Dependent Depletion (CADD Scores 1.4) method (PHRED scaled scores)^25^. The CADD algorithm includes conservation metrics, functional genomic data, exon-intron boundaries, and protein functionality scores. A score equal or greater of 20 indicates that a variant is predicted to be among the 1% of the most deleterious substitutions. We used a CADD score of over 23 to select for a damaging effect. In addition, we used a dbNSFP tool (database for nonsynonymous single nucleotide polymorphisms’ functional predictions) which incorporates five different algorithms (SIFT, Polyphen2, MutationTaster, MutationAssessor, and FATHMM)^26^. The most damaging variants were selected based on a voting system where four or all five of these algorithms predicted a negative functional impact.

## DISCUSSION

In this study, we employed whole-exome sequencing to human atherosclerotic tissue and can for the first time demonstrate the presence of somatic mutations as inherent characteristics of atherosclerotic plaques. Our findings suggest that the growth of atherosclerotic plaques does not solely result from random simultaneous division of numerous cells. Rather, certain mutation-carrying cells within the plaque tissue undergo local expansion, giving rise to clones consisting of large numbers of cells that span significant areas in various regions of the plaque and underlying media. This discovery not only sheds light on the cellular dynamics of atherosclerosis, but also places atherosclerotic lesions among a growing list of non-cancerous diseases where somatic mutations play a role, potentially influencing disease development^27^. Furthermore, our observations support the notion proposed over 50 years ago, suggesting that atherosclerotic plaques might indeed represent a form of neoplasia originating from vascular SMCs^6^.

To ensure the accurate identification of somatic mutations, each sample underwent two high-depth sequencing runs, and strict variant calling criteria were applied. The reported mutations’ reliability was substantiated through the identification of matching mutations in independently sequenced plaque samples from the same patients (and not from other patients). We also observed matching CHIP mutations in patient-matched buffy coats and plaque segments, providing additional validation. Additionally, the frequent occurrence of the same mutation in neighboring plaque samples indicates overlapping mutation-carrying clones between samples, further reinforcing the credibility of the identified mutations. Finally, the wide difference in numbers of identified mutations across patients, and the occurrence of mutation-negative plaque segments indicate a negligible false-positive rate.

One limitation of the analytical approach used in this study is its inability to fully assess the distribution of mutations across clonal populations within each plaque. This means that certain clones may contain numerous mutations, while other clones might harbor only a few mutations. In some cases, the total clone cell frequencies estimated from each mutation exceeded 100%, indicating that a scenario with a single mutation per clone is unlikely in these patients. To address this challenge and gain insights into the mutational architecture, we utilized a comparative approach.

Specifically, we identified groups of mutations with similar patterns, clustering mutations found in the same samples and displaying similar magnitudes and inter-sample differences in clone cell frequencies. This allowed us to suggest the presence of individual clones likely to carry multiple mutations using our bulk sequencing data. However, to establish the mutational architecture more definitively, more refined methodologies are necessary and should be pursued in future studies.

The design employed in this study centered on mutations identified within plaque tissue that remained undetectable in the corresponding blood samples of the same patients, enabling the identification of mutations specifically amplified within the plaque tissue. Although only a limited number of samples were investigated for the more detailed morphological location of mutations, we observed that most mutations were concentrated in the sub-core intimal layer, as opposed to the necrotic core, while the underlying medial layer exhibited relatively few mutations. This pattern may be due to the dominance of modulated SMCs in the sub-core intima, which originate from the medial layer and are known to be capable of migration and proliferation. These findings align with existing knowledge from mouse models, demonstrating that SMCs have the capacity to form clones also in experimental atherosclerosis^4,5^. In this context, it is particularly noteworthy that the enrichment of genes harboring missense or loss-of-function mutations, as opposed to synonymous mutations, prominently emphasized proteins related to the contractile apparatus (that are highly expressed in SMCs) and cell-cell adhesion. Disruption and misfolding of components of the contractile apparatus have been shown to lead to an atherosclerosis-promoting SMC phenotype^28^. Likewise, disruption of cell adhesion constitutes a pivotal component in the process of malignant transformation and the progression of cancer^29^. This phenomenon allows cells to detach from their natural microenvironment, relinquishing signals that maintain quiescence. Speculatively, the liberation of SMCs due to compromised cell-cell connections could mirror the phenomenon observed in cancer, where cells break free. Future research should investigate the consequence of identified mutations, and explore alternative experimental techniques and validation strategies to precisely elucidate the participation of SMCs or other cell types in the clonal expansion of mutated cells within atherosclerotic plaques. Furthermore, inquiries into potential associations with specific plaque subtypes and complications (including rupture, calcifications, and thrombosis) hold significant intrigue, yet obtaining answers necessitates a larger cohort of samples.

Regrettably, the current study is constrained by the relatively limited number of patients enrolled, impeding our capacity to thoroughly explore potential correlations between mutations and diverse clinical parameters, such as smoking habits, diabetes, medication utilization, and age. The scarcity of patient data available in extensive artery biobanks, complete with relevant sampled materials and comprehensive clinical profiles, poses a considerable challenge.

The central objective of this study was to demonstrate the presence of somatic mutations within human plaques, hereby establishing the existence of clonal cell populations. Despite the limitations imposed by our current sample size, we have successfully achieved this primary focus. Although our data indicate that the mutational spectrum is not random, it remains elusive to what extend somatic mutations contribute to clonal expansion. Indeed, as proposed for clonal expansion in mouse models^5,30^, clone precursors may be a subset of cells possessing advantageous properties related to *e.g.*, proliferation, survival in a toxic plaque environment, or escaping counter-regulatory mechanisms of the immune system. In line with this thinking, our mutated gene-enrichment results suggest that mutations occur in genes active in vascular smooth muscles. Alternatively, a clone-founding cell could merely be located “at the right place at the right time” thereby be the first to supersede a stimulatory threshold for activation, and as previously proposed, the selected clone may simultaneously suppress expansion of neighboring cells through juxtacrine lateral inhibition^5^. In this scenario, randomly amplified passenger mutations detected by whole-exome sequencing would only represent a fraction of clones. Finally, clonal patches carrying somatic mutations could already be present in the carotid artery prior to plaque development.

Another finding from our study pertains to the presence of CHIP-mutated hematopoietic clones in atherosclerotic plaque tissue. The most widespread understanding of how CHIP-carrying individuals face an elevated risk of atherosclerotic cardiovascular disease is that CHIP-mutations render leukocytes more pro-inflammatory, and that this in turn aggravates plaque formation^31^. However, this hypothesis requires direct evidence of CHIP-carrying cells being present in human atherosclerotic plaques, as we demonstrate in the current study.

In recent decades, the epidemiology of atherosclerotic cardiovascular disease has changed from being a disease of middle-aged smoking, hypercholesterolemic, hypertensive men, to increasingly being a disease of older people^32^.

This epidemiological transition is attributed to the successes of lipid- and blood pressure-lowering medication and smoking cessation initiatives, as well as a growing ageing population in most countries^32^. Consequently, the residual cardiovascular risk is increasingly attributed to age-related unknowns, which may show similarities with risk factors related to cancer, as suggested^33^. The identification of CHIP as an age-associated independent risk factor highlights somatic mutations and clonally expanded cells as significant contributors to atherosclerotic cardiovascular disease^7,8^. The present study demonstrates that similar processes take place locally in plaque tissues, but to what extent local somatic mutations and clonal expansion contribute to disease development and residual risk remains to beinvestigated. Interestingly, a recent exploratory analysis of the CANTOS trial showed that CHIP carriers with *TET2* mutations displayed significantly reduced relative risk (64%) of major cardiovascular events when treated with canakinumab (anti-IL1β antibody) as compared to non-CHIP carriers (reduced relative risk of 15%)^34^. This finding underscores the clinical relevance of understanding the biology of mutation-carrying clones in atherosclerotic cardiovascular disease. In the present study, we demonstrate that hematopoietic clones carrying mutations in *TET2* (and other CHIP genes) are not the only mutation-carrying clones that make up the plaque cell population. We envision that comprehending the biological underpinnings of clonal biology and phenotypic characteristics of expanding clones may unlock novel mechanisms to inhibit lesion development or increase plaque stability. To this end, another important notion from our study is that somatic mutations may serve as genetic tags for identification and characterization of clonal populations in future studies. This may prove to be a convenient tool for deciphering clonal biology in atherosclerosis to obtain unprecedented insight that could have implications for future therapeutics.

## ACKNOWLEDGEMENTS

We thank Mikkel Engstrøm Graversen, Jacob Fog Bentzon (Aarhus University Hospital, Aarhus, Denmark; and the Spanish National Center for Cardiovascular Research (CNIC), Madrid, Spain), and Iñigo Martincorena (Wellcome Sanger Institute, Cambridge, UK) for valuable discussion of the manuscript.

## DATA AVAILABILITY STATEMENT

The datasets generated and analyzed during the current study are not publicly available due to hospital guidelines and legislation regarding personal data. Data will be available from the corresponding author on reasonable request and with permission from Odense University Hospital Legal Department.

## REFERENCES

1. Diseases GBD, Injuries C. Global burden of 369 diseases and injuries in 204 countries and territories, 1990-2019: a systematic analysis for the Global Burden of Disease Study 2019. Lancet. 2020;396:1204–1222. doi: 10.1016/S0140-6736(20)30925-9

2. Bjorkegren JLM, Lusis AJ. Atherosclerosis: Recent developments. Cell. 2022;185:1630–1645. doi: 10.1016/j.cell.2022.04.004

3. Misra A, Rehan R, Lin A, Patel S, Fisher EA. Emerging Concepts of Vascular Cell Clonal Expansion in Atherosclerosis. Arterioscler Thromb Vasc Biol. 2022;42:e74–e84. doi: 10.1161/ATVBAHA.121.316093

4. Chappell J, Harman JL, Narasimhan VM, Yu H, Foote K, Simons BD, Bennett MR, Jorgensen HF. Extensive Proliferation of a Subset of Differentiated, yet Plastic, Medial Vascular Smooth Muscle Cells Contributes to Neointimal Formation in Mouse Injury and Atherosclerosis Models. Circ Res. 2016;119:1313–1323. doi: 10.1161/CIRCRESAHA.116.309799

5. Jacobsen K, Lund MB, Shim J, Gunnersen S, Fuchtbauer EM, Kjolby M, Carramolino L, Bentzon JF. Diverse cellular architecture of atherosclerotic plaque derives from clonal expansion of a few medial SMCs. JCI Insight. 2017;2. doi: 10.1172/jci.insight.95890

6. Benditt EP, Benditt JM. Evidence for a monoclonal origin of human atherosclerotic plaques. Proc Natl Acad Sci U S A. 1973;70:1753–1756. doi: 10.1073/pnas.70.6.1753

7. Jaiswal S, Fontanillas P, Flannick J, Manning A, Grauman PV, Mar BG, Lindsley RC, Mermel CH, Burtt N, Chavez A, et al. Age-related clonal hematopoiesis associated with adverse outcomes. N Engl J Med. 2014;371:2488–2498. doi: 10.1056/NEJMoa1408617

8. Jaiswal S, Natarajan P, Silver AJ, Gibson CJ, Bick AG, Shvartz E, McConkey M, Gupta N, Gabriel S, Ardissino D, et al. Clonal Hematopoiesis and Risk of Atherosclerotic Cardiovascular Disease. N Engl J Med. 2017;377:111–121. doi: 10.1056/NEJMoa1701719

9. Libby P, Ebert BL. CHIP (Clonal Hematopoiesis of Indeterminate Potential): Potent and Newly Recognized Contributor to Cardiovascular Risk. Circulation. 2018;138:666–668. doi: 10.1161/CIRCULATIONAHA.118.034392

10. Marnell CS, Bick A, Natarajan P. Clonal hematopoiesis of indeterminate potential (CHIP): Linking somatic mutations, hematopoiesis, chronic inflammation and cardiovascular disease. J Mol Cell Cardiol. 2021;161:98–105. doi: 10.1016/j.yjmcc.2021.07.004

11. Fuster JJ, MacLauchlan S, Zuriaga MA, Polackal MN, Ostriker AC, Chakraborty R, Wu CL, Sano S, Muralidharan S, Rius C, et al. Clonal hematopoiesis associated with TET2 deficiency accelerates atherosclerosis development in mice. Science. 2017;355:842–847. doi: 10.1126/science.aag1381

12. Lenormand C, Lipsker D. Somatic Mutations in “Benign” Disease. N Engl J Med. 2021;385:e34. doi: 10.1056/NEJMc2110545

13. Martincorena I, Roshan A, Gerstung M, Ellis P, Van Loo P, McLaren S, Wedge DC, Fullam A, Alexandrov LB, Tubio JM, et al. Tumor evolution. High burden and pervasive positive selection of somatic mutations in normal human skin. Science. 2015;348:880–886. doi: 10.1126/science.aaa6806

14. Grossmann S, Hooks Y, Wilson L, Moore L, O’Neill L, Martincorena I, Voet T, Stratton MR, Heer R, Campbell PJ. Development, maturation, and maintenance of human prostate inferred from somatic mutations. Cell Stem Cell. 2021;28:1262–1274 e1265. doi: 10.1016/j.stem.2021.02.005

15. Lawson ARJ, Abascal F, Coorens THH, Hooks Y, O’Neill L, Latimer C, Raine K, Sanders MA, Warren AY, Mahbubani KTA, et al. Extensive heterogeneity in somatic mutation and selection in the human bladder. Science. 2020;370:75–82. doi: 10.1126/science.aba8347

16. Martincorena I, Fowler JC, Wabik A, Lawson ARJ, Abascal F, Hall MWJ, Cagan A, Murai K, Mahbubani K, Stratton MR, et al. Somatic mutant clones colonize the human esophagus with age. Science. 2018;362:911–917. doi: 10.1126/science.aau3879

17. Ng SWK, Rouhani FJ, Brunner SF, Brzozowska N, Aitken SJ, Yang M, Abascal F, Moore L, Nikitopoulou E, Chappell L, et al. Convergent somatic mutations in metabolism genes in chronic liver disease. Nature. 2021;598:473–478. doi: 10.1038/s41586-021-03974-6

18. Olafsson S, McIntyre RE, Coorens T, Butler T, Jung H, Robinson PS, Lee-Six H, Sanders MA, Arestang K, Dawson C, et al. Somatic Evolution in Non-neoplastic IBD-Affected Colon. Cell. 2020;182:672–684 e611. doi: 10.1016/j.cell.2020.06.036

19. Anglesio MS, Papadopoulos N, Ayhan A, Nazeran TM, Noe M, Horlings HM, Lum A, Jones S, Senz J, Seckin T, et al. Cancer-Associated Mutations in Endometriosis without Cancer. N Engl J Med. 2017;376:1835–1848. doi: 10.1056/NEJMoa1614814

20. Cagan A, Baez-Ortega A, Brzozowska N, Abascal F, Coorens THH, Sanders MA, Lawson ARJ, Harvey LMR, Bhosle S, Jones D, et al. Somatic mutation rates scale with lifespan across mammals. Nature. 2022;604:517–524. doi: 10.1038/s41586-022-04618-z

21. Martinez-Jimenez F, Muinos F, Sentis I, Deu-Pons J, Reyes-Salazar I, Arnedo-Pac C, Mularoni L, Pich O, Bonet J, Kranas H, et al. A compendium of mutational cancer driver genes. Nat Rev Cancer. 2020;20:555–572. doi: 10.1038/s41568-020-0290-x

22. Wirka RC, Wagh D, Paik DT, Pjanic M, Nguyen T, Miller CL, Kundu R, Nagao M, Coller J, Koyano TK, et al. Atheroprotective roles of smooth muscle cell phenotypic modulation and the TCF21 disease gene as revealed by single-cell analysis. Nat Med. 2019;25:1280–1289. doi: 10.1038/s41591-019-0512-5

23. Pan H, Xue C, Auerbach BJ, Fan J, Bashore AC, Cui J, Yang DY, Trignano SB, Liu W, Shi J, et al. Single-Cell Genomics Reveals a Novel Cell State During Smooth Muscle Cell Phenotypic Switching and Potential Therapeutic Targets for Atherosclerosis in Mouse and Human. Circulation. 2020;142:2060–2075. doi: 10.1161/CIRCULATIONAHA.120.048378

24. Alsaigh T, Evans D, Frankel D, Torkamani A. Decoding the transcriptome of calcified atherosclerotic plaque at single-cell resolution. Commun Biol. 2022;5:1084. doi: 10.1038/s42003-022-04056-7

25. Rentzsch P, Witten D, Cooper GM, Shendure J, Kircher M. CADD: predicting the deleteriousness of variants throughout the human genome. Nucleic Acids Res. 2019;47:D886–D894. doi: 10.1093/nar/gky1016

26. Kircher M, Witten DM, Jain P, O’Roak BJ, Cooper GM, Shendure J. A general framework for estimating the relative pathogenicity of human genetic variants. Nat Genet. 2014;46:310–315. doi: 10.1038/ng.2892

27. Mustjoki S, Young NS. Somatic Mutations in “Benign” Disease. N Engl J Med. 2021;384:2039–2052. doi: 10.1056/NEJMra2101920

28. Kaw K, Chattopadhyay A, Guan P, Chen J, Majumder S, Duan XY, Ma S, Zhang C, Kwartler CS, Milewicz DM. Smooth muscle alpha-actin missense variant promotes atherosclerosis through modulation of intracellular cholesterol in smooth muscle cells. Eur Heart J. 2023;44:2713–2726. doi: 10.1093/eurheartj/ehad373

29. Janiszewska M, Primi MC, Izard T. Cell adhesion in cancer: Beyond the migration of single cells. J Biol Chem. 2020;295:2495–2505. doi: 10.1074/jbc.REV119.007759

30. Wang Y, Nanda V, Direnzo D, Ye J, Xiao S, Kojima Y, Howe KL, Jarr KU, Flores AM, Tsantilas P, et al. Clonally expanding smooth muscle cells promote atherosclerosis by escaping efferocytosis and activating the complement cascade. Proc Natl Acad Sci U S A. 2020;117:15818–15826. doi: 10.1073/pnas.2006348117

31. Jaiswal S, Ebert BL. Clonal hematopoiesis in human aging and disease. Science. 2019;366. doi: 10.1126/science.aan4673

32. Libby P. The changing landscape of atherosclerosis. Nature. 2021;592:524–533. doi: 10.1038/s41586-021-03392-8

33. Leiva O, AbdelHameid D, Connors JM, Cannon CP, Bhatt DL. Common Pathophysiology in Cancer, Atrial Fibrillation, Atherosclerosis, and Thrombosis: JACC: CardioOncology State-of-the-Art Review. JACC CardioOncol. 2021;3:619–634. doi: 10.1016/j.jaccao.2021.08.011

34. Svensson EC, Madar A, Campbell CD, He Y, Sultan M, Healey ML, Xu H, D’Aco K, Fernandez A, Wache-Mainier C, et al. TET2-Driven Clonal Hematopoiesis and Response to Canakinumab: An Exploratory Analysis of the CANTOS Randomized Clinical Trial. JAMA Cardiol. 2022;7:521–528. doi: 10.1001/jamacardio.2022.0386

